# Response of Unvaccinated US Adults to Official Information About the Pause in Use of the Johnson & Johnson-Janssen COVID-19 Vaccine

**DOI:** 10.1101/2021.06.08.21258558

**Authors:** Vishala Mishra, Joseph P. Dexter

## Abstract

On April 13, 2021 the Centers for Disease Control and Prevention (CDC) and Food and Drug Administration (FDA) recommended in a pause in use of the Johnson & Johnson (J&J)-Janssen COVID-19 vaccine due to reports of cerebral venous sinus thrombosis (CVST) in recently vaccinated individuals. The announcement of the pause required development of a coordinated communication strategy under extreme time pressure and careful messaging by stakeholders to mitigate reduced public confidence in COVID-19 vaccines, as was observed following the temporary suspension of the Oxford-AstraZeneca vaccine in many countries. In this survey study, we evaluated understanding and impressions of the CDC’s public online information about the J&J-Janssen pause among unvaccinated US adults.

## Introduction

On April 13, 2021 the Centers for Disease Control and Prevention (CDC) and Food and Drug Administration (FDA) recommended in a pause in use of the Johnson & Johnson (J&J)-Janssen COVID-19 vaccine due to reports of cerebral venous sinus thrombosis (CVST) in recently vaccinated individuals.^1^ The announcement of the pause required development of a coordinated communication strategy under extreme time pressure and careful messaging by stakeholders to mitigate reduced public confidence in COVID-19 vaccines, as was observed following the temporary suspension of the Oxford-AstraZeneca vaccine in many countries.^2,3^ In this survey study, we evaluated understanding and impressions of the CDC’s public online information about the J&J-Janssen pause among unvaccinated US adults.

## Methods

We administered the online survey to two cohorts of US adults recruited through Prolific between April 19-21, 2021 (cohort A) and April 21-23, 2021 (cohort B). The passages and survey questions are provided in eAppendix 1 and eAppendix 2. Cohort B was given an updated version of the web page text, reflecting changes made by the CDC on April 20, 2021.

Both cohorts were assembled using convenience sampling of unvaccinated adults; the first cohort was further restricted to individuals who expressed neutral or negative sentiments about COVID-19 vaccines (eMethods). The study was approved by Harvard University’s Committee on the Use of Human Subjects, and participants agreed to a consent statement on the first page of the survey.

Associations between participant characteristics and responses to the comprehension questions were assessed using ordinal logistic regression.

## Results

The Simple Measure of Gobbledygook readability grade was 12.9 for the first version and 12.7 for the second version of the passage (eMethods).

N = 271 and N = 286 participants were included in cohort A and cohort B, respectively; their demographic characteristics are listed in the Table. Across participants the median number of correct responses to the comprehension questions was 6 in both cohort A (IQR, 1.5; range, 0-7) and cohort B (IQR, 1.0; range, 1-7). In cohort B, a majority of respondents answered that a person who develops a severe headache after receiving the J&J-Janssen vaccine should self-monitor for 24 hours (87 [30.4%]) or report the side effect through CDC’s v-safe application (74 [25.9%]) instead of seeking immediate medical care for possible CVST (eTable1). Total number of correct responses was negatively associated with intention not to seek vaccination in both cohort A (odds ratio [OR], 0.61; 95% CI, 0.45-0.82; *P* = .001; eTable2) and cohort B (OR, 0.48; 95% CI, 0.31-0.74; *P* = .001; eTable3).

**Table.** **Characteristics and Responses of Participants Who Completed the Online Surveys About the J&J-Janssen Vaccine Pause**

|  | Cohort A (N = 271) | Cohort B (N = 286) |
| --- | --- | --- |
| Characteristic | No. (%) | No. (%) |
| Age, y |  |  |
| 18-29 | 119 (43.9) | 145 (50.7) |
| 30-49 | 116 (42.8) | 111 (38.9) |
| ≥50 | 36 (13.3) | 30 (10.5) |
| Gender |  |  |
| Female | 144 (53.1) | 136 (47.6) |
| Male | 124 (45.8) | 144 (50.3) |
| Non-binary, transgender, or other | 3 (1.1) | 6 (2.1) |
| Ethnicity |  |  |
| Hispanic or Latinx | 54 (20.0) | 39 (13.6) |
| Race<sup>a</sup> |  |  |
| Asian | 29 (10.7) | 96 (33.6) |
| Black or African American | 70 (25.8) | 37 (12.9) |
| White | 160 (59.0) | 139 (48.6) |
| Other<sup>b</sup> | 27 (10.0) | 28 (9.8) |
| Educational attainment |  |  |
| Less than high school diploma | 3 (1.1) | 6 (2.1) |
| High school diploma or equivalent | 54 (19.9) | 46 (16.1) |
| Some college or associate's degree | 102 (37.6) | 106 (37.1) |
| Bachelor's degree | 81 (29.9) | 102 (35.7) |
| Graduate or professional degree | 31 (11.4) | 26 (9.1) |
| Political partisanship |  |  |
| Democratic (including leaners) | 81 (29.9) | 150 (52.4) |
| Republican (including leaners) | 91 (33.6) | 42 (14.7) |
| Independent or other | 99 (36.5) | 94 (32.9) |
| Geography |  |  |
| Rural area | 41 (15.1) | 35 (12.2) |
| Suburban or urban area | 230 (84.9) | 251 (87.8) |
| Intention to receive vaccine |  |  |
| Definitely will not | 71 (26.2) | 29 (10.1) |
| Probably will not | 68 (25.1) | 17 (5.9) |
| Undecided | 54 (19.9) | 29 (10.1) |
| Definitely or probably will | 78 (28.8) | 211 (73.8) |
| Comprehension questions<sup>c</sup> |  |  |
| Reason for pause | 259 (95.6) | 280 (97.9) |
| Causal relationship between vaccine and side effect | 196 (72.3) | 215 (75.2) |
| Affected population | 234 (86.3) | 248 (86.7) |
| Safety of mRNA vaccines | 214 (79.0) | 250 (87.4) |
| Revaccination of J&J-Janssen recipients | 164 (60.5) | 189 (66.1) |
| Symptom monitoring for J&J-Janssen recipients | 180 (66.4) | 121 (42.3) |
| Rescheduling of cancelled appointments | 192 (70.8) | 229 (80.1) |
| Comprehension score<sup>d</sup> |  |  |
| 7 | 62 (22.9) | 43 (15.0) |
| 6 | 80 (29.5) | 119 (41.6) |
| 5 | 61 (22.5) | 59 (20.6) |
| 4 | 33 (12.2) | 38 (13.3) |
| 3 | 24 (8.9) | 18 (6.3) |
| 0-2 | 11 (4.1) | 9 (3.1) |
| Self-reported impressions of passage<sup>e</sup> |  |  |
| Accurate and should be trusted | 119 (43.9) | 198 (69.2) |
| High-quality evidence | 110 (40.6) | 180 (62.9) |
| Clear and easy to read | 229 (84.5) | 243 (85.0) |
| Understandable | 229 (84.5) | 251 (87.8) |
| Requiring great effort to understand | 58 (21.4) | 69 (24.1) |
| Other people would want to read | 173 (63.8) | 172 (60.1) |
| Would share on social media | 53 (19.6) | 75 (26.2) |

<sup>a</sup>Participants could select more than one option.
<sup>b</sup>Includes participants who selected "American Indian or Alaska Native," "Native Hawaiian or Other Pacific Islander," or "Another option not listed here."
<sup>c</sup>Number of participants who gave the correct answer to each question.
<sup>d</sup>Number of participants who gave the indicated number of correct answers across all questions.
<sup>e</sup>Number of participants who answered "Strongly agree" or "Agree" about each description on a 6-point Likert scale.

The web page mentioned “a small number of reports” of CVST in individuals who received the J&J-Janssen vaccine. When asked to guess a specific number, 188 (69.4%) respondents in cohort A and 133 (46.5%) respondents in cohort B estimated 100 or more cases, at least an order of magnitude higher than the actual value; 176 (64.9%) respondents in cohort A and 128 (44.8%) respondents in cohort B estimated 10 or more deaths after vaccination (Figure). In addition, over one third of participants in both cohorts (126 [46.5%] and 106 [37.1%], respectively) predicted that the pause would last longer than 2 months.

**Figure.**
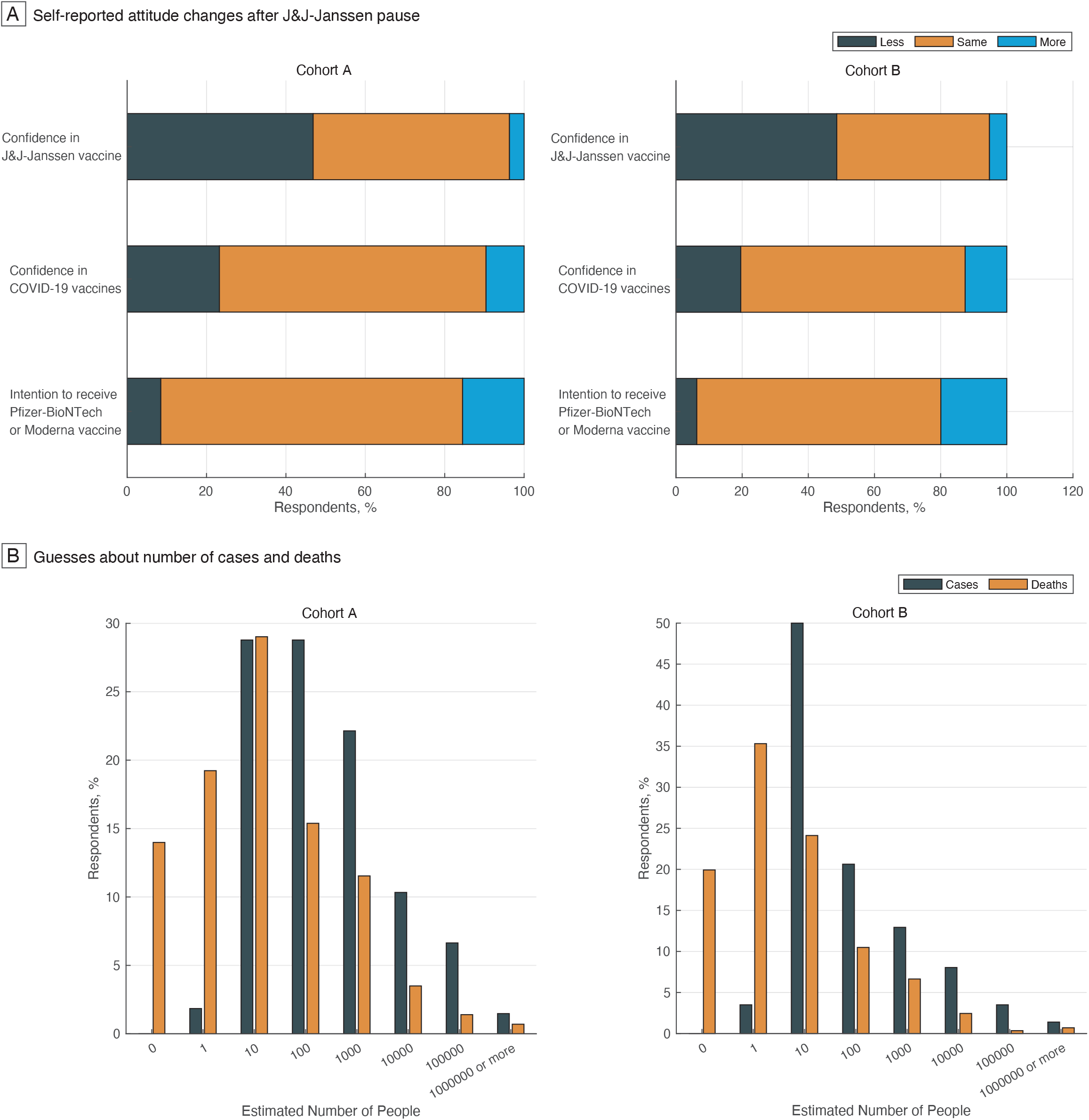
Self-Reported Attitude Changes and Estimated Cases and Deaths. The graphs show (A) the number of respondents who expressed changes in attitude before and after the pause in response to three counterfactual questions and (B) the estimated number of cases and deaths that led to the pause. 0 was not included as an answer choice for the question about cases.

In response to a counterfactual question (eMethods), 127 (46.9%) participants in cohort A and 139 (48.6%) participants in cohort B indicated that the pause reduced their confidence in the safety of the J&J-Janssen vaccine (Figure). A majority of participants reported no change in their confidence in the safety of COVID-19 vaccines in general (182 [67.2%] for cohort A, 194 [67.8%] for cohort B) or intention to receive the Pfizer-BioNTech or Moderna vaccine (206 [76.0%] for cohort A, 211 [73.8%] for cohort B).

## Discussion

After reading online information from the CDC about the J&J-Janssen pause, many respondents overestimated the number of case reports that prompted the pause. Since verbal descriptors are elastic concepts that can be misinterpreted, grounding them with numbers can reduce variability in risk perception and promote informed decision making.^4^

Respondents also expressed reduced confidence in the safety of the J&J-Janssen vaccine, highlighting the danger of conveying piecemeal information about risk during a pandemic response.^3^ The negative association between understanding of the passage and self-reported vaccine hesitancy, and the unwillingness of most participants to share the information on social media, suggest a need for more targeted and accessible messaging to promote confidence in the J&J-Janssen vaccine among unvaccinated adults.^5,6^

The study is limited by the convenience sampling strategy, which was used to investigate response to the J&J-Janssen vaccine pause soon after it was announced. The participants recruited were not representative of the US population as a whole, and the findings of this study should not be generalized to other contexts.

## Supporting information

Supplemental Online Content

## Data Availability

Raw data is available from the corresponding author on request.

## Author Contributions

Drs Mishra and Dexter had full access to all of the data in the study and take responsibility for the integrity of the data and the accuracy of the data analysis.

### Concept and design

Both authors.

### Acquisition, analysis, or interpretation of data

Both authors.

### Drafting of the manuscript

Both authors.

### Critical revision of the manuscript for important intellectual content

Both authors.

### Statistical analysis

Both authors.

### Obtained funding

Dexter.

### Supervision

Both authors.

## References

1. Karron RA, Key NS, Sharfstein JM. Assessing a rare and serious adverse event following administration of the Ad26.COV2.S vaccine. JAMA. Published online April 30, 2021. doi:10.1001/jama.2021.7637

2. Scott M. Trust in AstraZeneca vaccine wanes across EU, survey finds. Published March 22, 2021. Accessed June 6, 2021. https://www.politico.eu/article/trust-oxford-astrazeneca-coronavirus-vaccine-wanes-europe-survey/

3. Wood S, Schulman K. Beyond politics—Promoting Covid-19 vaccination in the United States. N Engl J Med. 2021;384(7):e23. doi:10.1056/NEJMms2033790

4. Edwards A, Elwyn G, Mulley A. Explaining risks: turning numerical data into meaningful pictures. BMJ. 2002;324(7341):827–830. doi:10.1136/bmj.324.7341.827

5. Hamel L, Lopes L, Kearney A, Brodie M. KFF COVID-19 vaccine monitor: March 2021. Published March 30, 2021. Accessed June 6, 2021. https://www.kff.org/coronavirus-covid-19/poll-finding/kff-covid-19-vaccine-monitor-march-2021/

6. Mishra V, Dexter JP. Comparison of readability of official public health information about COVID-19 on websites of international agencies and the governments of 15 countries. JAMA Netw Open. 2020;3(8):e2018033. doi:10.1001/jamanetworkopen.2020.1803

