## Supplemental Online Content for "Response of Unvaccinated US Adults to Official Information About the Pause in Use of the Johnson & Johnson-Janssen COVID-19 Vaccine"

**eMethods**

**eAppendix 1.** Survey Administered to Cohort A

**eAppendix 2.** Survey Administered to Cohort B

**eTable 1.** Full Responses to Comprehension Questions

**eTable 2.** Summary of Ordinal Logistic Regression Analysis for Cohort A

**eTable 3.** Summary of Ordinal Logistic Regression Analysis for Cohort B

**eReferences**

This supplemental material has been provided by the authors to give readers additional information about their work.

**eMethods**

*Demographic Information*

Participants self-reported demographic information about age, gender, ethnicity, race, educational attainment, political partisanship and ideology, and place of residence. Information about race and ethnicity was collected because of differing rates of COVID-19 vaccination across racial and ethnic minority groups.[1,2]

*Assessment of COVID-19 Vaccine Hesitancy*

The survey included two questions about COVID-19 vaccine hesitancy adapted from Persad *et al*. 2021,[3] which were presented before the passage. The first question asked "How likely are you to get vaccinated for COVID-19 when a vaccine is available for you?" and included 6 answer choices ("I definitely will NOT get vaccinated."; "I probably will NOT get vaccinated."; "I probably will get vaccinated."; "I definitely will get vaccinated."; "I am completely undecided about whether I will get vaccinated."; "I have already been vaccinated.") The second question asked "Thinking about this in a different way, which of the following statements comes closest to what you are most likely to do when a COVID-19 vaccine is available for you?" and included 4 answer choices ("I will get vaccinated as soon as possible."; "I will wait to see what happens with other people before deciding whether to get vaccinated myself."; "I will not get vaccinated, regardless of what happens to other people who get the vaccine."; "I have already been vaccinated.")

*Prescreening and Sampling Strategy*

We administered the surveys through Prolific (https://www.prolific.co/), an online survey vendor with a large number of active users in the United States. Both cohorts were assembled using a convenience sampling strategy in which participants enrolled on a first-come, first-serve basis. Prescreening questions written by Prolific and previously administered to their users were employed to restrict enrollment to the target populations.

The following prescreening criteria were applied to both cohorts:

Question: "In which country do you currently reside?"

Accepted answer: "United States"

Question: "Have you received a coronavirus (COVID-19) vaccination?"

Accepted answer: “No”

One additional criterion regarding vaccine hesitancy was applied only to cohort A:

Question: “Please describe your attitudes towards the COVID-19 (Coronavirus) vaccines:”

Accepted answers: “Against (I feel negatively about the vaccines)”; “Neutral (I don’t have strong opinions either way)”

In addition, participants in related pilot studies run by the investigators between February 1, 2021 and April 15, 2021 were excluded during prescreening, and all participants in cohort A were ineligible to join cohort B.

To increase the racial and ethnic diversity of the convenience samples, we limited the number of participants who could enroll in each cohort based on their answers to the following prescreening question:[4]

“Please indicate your ethnicity (i.e. peoples’ ethnicity describes their feeling of belonging and attachment to a distinct group of a larger population that shares their ancestry, colour, language or religion)?”

For each cohort, we capped enrollment at a maximum of 100 people for each of the following 5 groups:

Group 1: Responded “White/Caucasian”
Group 2: Responded “Black/African American”
Group 3: Responded “East Asian”, “South Asian”, or “South East Asian”
Group 4: Responded “African”, “Caribbean”, “Middle Eastern”, “Mixed”, “Native American or Alaskan Native”, “Other (please feel free to us know your ethnicity via email)”, “White/Sephardic Jew”, “Black/British”, “White Mexican”, or “Romani/Traveller”
Group 5: Responded “Latino/Hispanic”

*Participant Compensation*

Participants were paid $2.00 for taking the survey.

*Exclusion Criteria*

303 participants enrolled in cohort A, and 286 participants enrolled in cohort B. After data collection, we excluded participants who met any of the following criteria:

1) Did not complete the whole survey.

2) Answered “Not at all carefully” to the question “How carefully did you complete this survey? Please answer honestly. Your payment does NOT depend on your response to this question.”

3) Gave incorrect responses to both attention check questions, “Please answer ‘Slightly unlikely’ to this question.” and “What color is the sky? Please answer this question incorrectly, on purpose, by choosing ‘Red’ instead of ‘Blue.’ ”

4) Answered “Yes, one dose” or “Yes, two doses” to the question “Have you gotten a COVID-19 vaccine?”

In addition, participants were excluded from cohort A if they reported a strong intention to receive a COVID-19 vaccine, in contrast to their answers to the Prolific prescreening questions. For these purposes, strong intention was defined as answering “I definitely will get vaccinated” to the question “How likely are you to get vaccinated for COVID-19 when a vaccine is available for you?” and answering “I will get vaccinated as soon as possible” to the question “Thinking about this in a different way, which of the following statements comes closest to what you are most likely to do when a COVID-19 vaccine is available for you?”

Participants who reported using Google searches or other outside help to answer the survey questions (9 [3.3%] in cohort A and 10 [3.5%] in cohort B) were not excluded.

*CDC Passages and Readability Analysis*

The survey passages were taken from https://www.cdc.gov/coronavirus/2019-ncov/vaccines/safety/JJUpdate.html, which is the CDC’s primary web page for communicating information about the pause to the general public. To create the text included in the survey, we removed links and references to other web pages, non-textual elements such as figures and infographics, and formatting other than section headers, bulleted lists, and paragraph breaks. No other changes were made to the passages.

Participants in cohort A were presented with the version of the passage released on April 16, 2021. It is 948 words long and scores above an eighth grade reading level by both the Simple Measure of Gobbledygook (SMOG) and Flesch-Kincaid Grade Level (FKGL) metrics (12.9 and 11.4, respectively).

On April 20, 2021 the CDC made several changes to the passage, including reducing the length to 795 words and altering the language and section headings. The core content was unaltered.

Participants in cohort B were given the revised passage, which scores 12.7 by SMOG and 10.7 by FKGL.

The full text of each passage is available in eAppendix 1 and eAppendix 2, respectively. Readability grade levels were calculated using Readability Studio Professional, version 2020 (Oleander Software).

*Comprehension Questions*

We developed 7 multiple-choice questions to assess how participants understood the major recommendations of the passage. The full set of questions and answer choices, along with participant responses, are provided in eTable 1. The first 3 questions involve recall of information stated directly in the passage; the remaining 4 questions involve application of concepts to hypothetical scenarios.

*Impression Questions*

Participants were asked to rate their impressions of 7 attributes of the passage using a 6-point Likert scale (options “Strongly disagree”, “Disagree”, “Slightly disagree”, “Slightly agree”, “Agree”, and “Strongly agree”). The questions were adapted from a previous study of COVID-19 health literacy by Kerr *et al*. 2021.[5]

*Counterfactual Questions*

We used the nonrandomized counterfactual format described Graham and Coppock 2021 to assess participants’ beliefs about the effect of the J&J/Janssen pause on their confidence in the safety of COVID-19 vaccines and the likelihood that they would receive the Pfizer/BioNTech or Moderna vaccine.[6] Participants first answered three questions about their current beliefs after reading the passage. They were then instructed to answer the same three questions as if they had been asked before the pause was announced.[6,7] For the figure, we tabulated the number of participants who expressed more, less, or the same confidence in response to the counterfactual questions than the baseline questions.

*Statistical analysis*

Associations between participant characteristics and performance on the comprehension questions were assessed using ordinal logistic regression. The dependent variable was the total number of correct responses to the 7 comprehension questions. Statistical analyses were performed using the Python package statsmodels (version 0.13.0.dev0). Statistical significance was defined as $P < .05$.

**eAppdenix 1. Survey Administered to Cohort A**

*Please answer the following questions about your experiences with the COVID-19 pandemic and COVID-19 vaccines.*

Around how often do you look for information about COVID-19 on the internet?

Never
Once a month
2-3 times per month
2-4 Once a week
Multiple times per week
Once a day
Multiple times per day

Do you ever share information about COVID-19 with your social media network?

No
Yes
I do not have an active account on a social media platform.

Which cable news network do you watch most often?

CNN
Fox News
International news channels (such as BBC, Al Jazeera, or Euronews)
MSNBC
I do not watch cable news.

As far as you know, do you have COVID-19 now, or have you had COVID-19 in the past?

No
Yes
I don't know.

As far as you know, has anyone in your immediate social circle, such as close friends or family members, had COVID-19?

No
Yes
I don't know.

Are you currently eligible to get a COVID-19 vaccine where you live?

No
Yes
I don't know.

Have you gotten a COVID-19 vaccine?

No

Yes, one dose

Yes, two doses

How likely are you to get vaccinated for COVID-19 when a vaccine is available for you?

I definitely will NOT get vaccinated.

I probably will NOT get vaccinated.

I probably will get vaccinated.

I definitely will get vaccinated.

I am completely undecided about whether I will get vaccinated.

I have already been vaccinated.

Thinking about this in a different way, which of the following statements comes closest to what you are most likely to do when a COVID-19 vaccine is available for you?

I will get vaccinated as soon as possible.

I will wait to see what happens with other people before deciding whether to get vaccinated myself.

I will not get vaccinated, regardless of what happens to other people who get the vaccine.

I have already been vaccinated.

Have you scheduled a COVID-19 vaccine appointment?

Yes, I already got a COVID-19 vaccine.

Yes, I have an upcoming COVID-19 vaccine appointment.

Yes, but my COVID-19 vaccine appointment was cancelled.

No, I don't know how to schedule a COVID-19 vaccine appointment.

No, I am not yet eligible to get a COVID-19 vaccine.

No, I don't plan to schedule a COVID-19 vaccine appointment.

Below are some reasons people may give for why they will NOT get a COVID-19 vaccine. For each one, please indicate whether it definitely applies to you, somewhat applies to you, or does not apply to you as a reason for not getting a COVID-19 vaccine.

| | Definitely applies to me | Somewhat applies to me | Does not apply to me |
|---|---|---|---|
| I have a health condition that would make it risky for me to get a vaccine. | ○ | ○ | ○ |
| I have a right not to get vaccinated and choose to exercise that right. | ○ | ○ | ○ |
| I do not trust vaccines in general. | ○ | ○ | ○ |
| I do not think the vaccine will be effective in protecting me from COVID-19. | ○ | ○ | ○ |
| I do not think the vaccine will be safe because it could have harmful side effects. | ○ | ○ | ○ |
| I am concerned approval of the vaccine has been rushed, without adequate testing for safety and effectiveness. | ○ | ○ | ○ |
| I do not think it is necessary to get vaccinated because COVID-19 is not as big of a problem as it is being made out to be. | ○ | ○ | ○ |
| I do not think it is necessary to get vaccinated because I take precautions to protect myself, such as wearing a mask, maintaining physical distance, and washing hands often. | ○ | ○ | ○ |
| I do not think I will get very sick if I get COVID-19. | ○ | ○ | ○ |
| I am not able to get a COVID-19 vaccine appointment. | ○ | ○ | ○ |

*Please answer the following two questions just to show that you're paying attention to the survey.*

Please answer “Slightly unlikely” to this question.

- Very unlikely
- Unlikely
- Slightly unlikely
- Slightly likely
- Likely
- Very likely

What color is the sky? Please answer this question incorrectly, on purpose, by choosing “Red” instead of “Blue.”

- Blue
- Green
- Red
- Yellow

*Please read the following text passage about the J&J/Janssen COVID-19 vaccine. After you read the passage, you will be asked some questions. You may take as much time as you would like to read the passage, and you may look back at the passage as you answer the questions.*

***

**Recommendation to Pause Use of Johnson & Johnson’s Janssen COVID-19 Vaccine**

On April 13, 2021, CDC and FDA recommended a pause in the use of Johnson & Johnson’s Janssen COVID-19 Vaccine. Of the nearly 7 million doses administered so far in the United States, a small number of reports of a rare and severe type of blood clot have been reported in people after receiving the J&J/Janssen COVID-19 Vaccine. All reports occurred among women between the ages of 18 and 48, and symptoms occurred six to 13 days after vaccination. As of April 13, 2021, of the more than 180 million doses administered so far of the Pfizer-BioNTech or Moderna vaccines, no reports matching those associated with the J&J/Janssen vaccine have been received.

**J&J/Janssen COVID-19 Vaccine Update, April 13, 2021**

The use of this vaccine is ‘paused’ for now. This is because the safety systems that make sure vaccines are safe received a small number of reports of a rare and severe type of blood clot happening in people who got this vaccine.

We do not know enough yet to say if the vaccine is related to or caused this health issue. To be extra careful, CDC and FDA recommend that the vaccine not be given until we learn more.

If you got this vaccine, seek medical care urgently if you develop any of the following symptoms:

- severe headache,
- backache,
- new neurologic symptoms,
- severe abdominal pain,
- shortness of breath,
- leg swelling,
- tiny red spots on the skin (petechiae),
- or new or easy bruising

**Learn More About the J&J/Janssen Vaccine Pause**

**What does a "pause" mean?**

On April 13, 2021, CDC and the US Food and Drug Administration (FDA) recommended a pause in the use of Johnson & Johnson's Janssen (J&J/Janssen) COVID-19 Vaccine. Although the J&J/Janssen vaccine is still authorized for use, CDC and FDA recommend this vaccine not be given to anyone until we know more. This gives scientists a chance to review the data and decide if recommendations on who should get the vaccine need to change. CDC and FDA will share more information as soon as possible with healthcare providers, people who got the vaccine, and the public.

**What do I need to know about the possible safety issue?**

Here is what we know now: scientists and doctors always look carefully at all reported side effects.

From their review, they saw a small number of cases of a rare and severe type of blood clot in people who got the J&J/Janssen COVID-19 Vaccine. All reported cases were in women between the ages of 18 and 48, and the problems were found up to two weeks after vaccination.

**What if I got this vaccine?**

If you got this vaccine more than three weeks ago, your risk of developing a blood clot is very low.

If you got this vaccine within the last three weeks, your risk of developing a blood clot is also very low. However, you should be on the lookout for possible symptoms of a blood clot:

- Severe headache
- Blurred vision
- Fainting
- Seizures
- Pain in your abdomen (chest or stomach)
- Leg pain or swelling
- Shortness of breath

Get medical care right away if you have any of these symptoms and got the J&J/Janssen COVID-19 Vaccine within the last few weeks. If you have any questions at all, call your doctor, nurse, or clinic.

**Has this issue been seen with the other COVID-19 vaccines?**

No. As of April 13, 2021, no cases of this blood clot issue have been reported among the more than 180 million people who received the Pfizer-BioNTech or Moderna vaccines.

**Are COVID-19 vaccines safe?**

Yes. COVID-19 vaccine safety is a top priority for the federal government, and all reports of health problems following COVID-19 vaccination are taken very seriously and investigated as needed. We know the safety systems in place are working. COVID-19 vaccines have undergone and will continue to undergo the most intensive safety monitoring in U.S. history.

**Should I cancel my vaccination appointment?**

If you are scheduled to get the J&J/Janssen COVID-19 Vaccine, work with your vaccine provider to reschedule your appointment to get another type of COVID-19 vaccine.

**Safety Is a Top Priority**

COVID-19 vaccine safety is a top priority for the federal government, and all reports of health problems following COVID-19 vaccination are taken very seriously. This potential safety issue was caught early, and this pause reflects the federal government’s commitment to transparency as CDC and FDA review these data. COVID-19 vaccines have undergone and will continue to undergo the most intensive safety monitoring in U.S. history.

**What to Do If You Received the J&J/Janssen COVID-19 Vaccine**

If you received the vaccine more than three weeks ago, the risk of developing a blood clot is likely very low at this time.

If you received the vaccine within the last three weeks, your risk of developing a blood clot is also very low and that risk will decrease over time.

Contact your healthcare provider and seek medical treatment urgently if you develop any of the following symptoms:

- severe headache,
- backache,
- new neurologic symptoms,
- severe abdominal pain,
- shortness of breath,
- leg swelling,
- tiny red spots on the skin (petechiae), or
- new or easy bruising

If you are scheduled to get the J&J/Janssen COVID-19 Vaccine, please work with your vaccine provider to reschedule your appointment to receive another authorized and recommended COVID-19 vaccine. There are two other COVID-19 vaccines authorized and recommended for use in the United States: Pfizer-BioNTech and Moderna.

If you experience any adverse events after vaccination, report them to v-safe and the Vaccine Adverse Event Reporting System.

***

Thinking about the information you just read, how confident are you in the safety of COVID-19 vaccines in general?

Not confident at all
Slightly confident
Somewhat confident
Fairly confident
Completely confident

Thinking about the information you just read, how confident are you in the safety of the J&J/Janssen COVID-19 vaccine in particular?

Not confident at all
Slightly confident
Somewhat confident
Fairly confident
Completely confident

Thinking about the information you just read, how likely would you be to get either the Moderna or Pfizer/BioNTech COVID-19 vaccine?

I definitely would NOT get either the Moderna or Pfizer/BioNTech vaccine.
I probably would NOT get either the Moderna or Pfizer/BioNTech vaccine.
I probably would get either the Moderna or Pfizer/BioNTech vaccine.
I definitely would get either the Moderna or Pfizer/BioNTech vaccine.
I am completely undecided about whether I would get either the Moderna or Pfizer/BioNTech vaccine.
I have already gotten either the Moderna or Pfizer/BioNTech vaccine.
I have already gotten a different COVID-19 vaccine.

*Please answer the same three questions as if we had asked you BEFORE the pause of the J&J/Janssen vaccine was announced.*

Now imagine that we had asked you these three questions BEFORE the pause of the J&J/Janssen vaccine was announced. How would you have answered the first question, which is how confident are you in the safety of COVID-19 vaccines in general?

Not confident at all
Slightly confident
Somewhat confident
Fairly confident
Completely confident

Now imagine that we had asked you these three questions BEFORE the pause of the J&J/Janssen vaccine was announced. How would you have answered the second question, which is how confident are you in the safety of the J&J/Janssen COVID-19 vaccine in particular?

Not confident at all
Slightly confident
Somewhat confident
Fairly confident
Completely confident

Now imagine that we had asked you these three questions BEFORE the pause of the J&J/Janssen vaccine was announced. How would you have answered the third question, which is how likely would you be to get either the Moderna or Pfizer/BioNTech COVID-19 vaccine if available to you?

I definitely would NOT get either the Moderna or Pfizer/BioNTech vaccine.
I probably would NOT get either the Moderna or Pfizer/BioNTech vaccine.
I probably would get either the Moderna or Pfizer/BioNTech vaccine.
I definitely would get either the Moderna or Pfizer/BioNTech vaccine.
I am completely undecided about whether I would get either the Moderna or Pfizer/BioNTech vaccine.
I have already gotten either the Moderna or Pfizer/BioNTech vaccine.
I have already gotten a different COVID-19 vaccine.

*Using a six-point scale, please indicate how strongly you agree with each of the following statements about the passage.*

I think that the information in the passage is accurate and should be trusted.

Strongly disagree
Disagree
Slightly disagree
Slightly agree
Agree
Strongly agree

I think that the information in the passage is based on high-quality evidence.

Strongly disagree
Disagree
Slightly disagree
Slightly agree
Agree
Strongly agree

I think that the writing in the passage is clear and easy to read.

Strongly disagree
Disagree
Slightly disagree
Slightly agree
Agree
Strongly agree

I think that I understand the information in the passage.

Strongly disagree
Disagree
Slightly disagree
Slightly agree
Agree
Strongly agree

I think that I had to put a lot of effort into understanding the information in the passage.

Strongly disagree
Disagree
Slightly disagree
Slightly agree
Agree
Strongly agree

I think that other people would want to read the passage.

Strongly disagree
Disagree
Slightly disagree
Slightly agree
Agree
Strongly agree

I think that I would share the information in the passage with friends on social media.

Strongly disagree
Disagree
Slightly disagree
Slightly agree
Agree
Strongly agree

*Please answer the following questions with your best guess.*

As you read in the passage, almost 7 million people have received the J&J/Janssen vaccine in the U.S. *If you had to guess*, about how many of them have developed a rare and severe type of blood clot after being vaccinated?

1 person
10 people
100 people

1,000 people
10,000 people
100,000 people
1,000,000 or more people

As you read in the passage, almost 7 million people have received the J&J/Janssen vaccine in the U.S. *If you had to guess*, about how many of them have died from a rare and severe type of blood clot after being vaccinated?

0 people
1 person
10 people
100 people
1,000 people
10,000 people
100,000 people
1,000,000 or more people

*If you had to guess*, how much longer do you think the pause in use of the J&J/Janssen COVID-19 vaccine will last?

1 day
2-6 days
1-2 weeks
2-4 weeks
1-2 months
More than 2 months (but not permanent)
I think the pause will be permanent.

*Thank you for reading the text passage about the J&J/Janssen COVID-19 vaccine and answering our questions.*

*Now please answer a second set of questions based only on the information in the same passage that you just read. As before, you may take as much time as you would like to reread the passage, and you may look back at the passage as you answer the questions.*

Is it currently legal to give the J&J/Janssen vaccine in the U.S.?

No, because the FDA has removed its emergency authorization.
No, because its use has been paused by the FDA and CDC.
Yes, because the pause in use is a recommendation, not a requirement.
The passage does not say.

Why are the CDC and FDA recommending a pause in use of the J&J/Janssen vaccine?

After being approved for emergency use in the U.S., the vaccine was found to be ineffective at preventing COVID-19 infections, hospitalizations, and deaths.

Problems were recently identified with how the original clinical trial was run.

J&J has had trouble making the vaccine, and almost no doses are available now.

A possible safety issue has been identified with the vaccine, and more time is needed to study the issue.

Does the J&J/Janssen COVID-19 vaccine cause blood clots?

No

Yes

More information is needed to know for sure.

A rare and severe type of blood clot has been reported in ________ who received the J&J/Janssen vaccine.

women

men

both men and women

people over the age of 50

On April 1st John got his first shot of the Pfizer/BioNTech vaccine. The day after getting vaccinated, he felt tired and had a mild headache. His second shot is scheduled for this week. Should John cancel the appointment because of safety concerns?

No, because a rare and severe type of blood clot has only been reported in people who received the J&J/Janssen vaccine, not people who received the Moderna or Pfizer/BioNTech vaccines.

No, because blood clots are only caused by the first dose of Pfizer/BioNTech vaccine, not the second.

Yes, because he had a headache after the first dose.

Yes, because possible safety issues with the J&J/Janssen vaccine might apply to all COVID-19 vaccines.

Elizabeth got the the J&J/Janssen COVID-19 six weeks ago. Since then she has not had any side effects from the vaccine or symptoms of COVID-19. In light of the pause, she should ________.

get the Moderna vaccine

get the Pfizer/BioNTech vaccine

ask her doctor for advice about getting a different vaccine

do nothing different

Linda gets the J&J/Janssen vaccine on Monday. On Friday she develops a severe headache. What should she do first?

Seek urgent medical care for a possible blood clot.
Monitor herself for 24 hours and then seek medical care if the headache has not improved.
Do nothing, because headaches are a normal side effect of COVID-19 vaccines.
Report the symptom through v-safe.

Jessie's appointment to get the J&J/Janssen vaccine has been canceled because of the pause. What should they do now?

Work with their vaccine provider to reschedule the appointment and get a different COVID-19 vaccine.
Wait to get vaccinated until the J&J/Janssen vaccine is available again.
Show up for the canceled appointment and ask to get a different vaccine.
Either the first or the second answer is correct.

*Please answer the following questions about your background.*

How old are you?

18-29
30-39
40-49
50-64
65 or older

Which best describes your gender? Please choose as many options as apply.

Female
Male
Non-binary
Transgender
Another option not listed here (please specify): ______________________________

Are you Hispanic or Latino/Latina/Latinx?

Not Hispanic or Latino/Latina/Latinx
Hispanic or Latino/Latina/Latinx

Which best describes your race? Please choose as many options as apply.

American Indian or Alaska Native
Asian

Black or African American
Native Hawaiian or Other Pacific Islander
White
Another option not listed here (please specify): ______________________________________________

What is your highest level of formal education?
Some high school or less
High school diploma or equivalent
Some college or associate's degree
Bachelor's degree
Graduate or professional degree

Generally speaking, which of the following best describes your political affiliation?
Strong Republican
Leaning Republican
Leaning Democratic
Strong Democratic
Independent
Another option not listed here (please specify): ______________________________________________

Which candidate did you vote for in the 2020 U.S. presidential election?
Joe Biden (Democratic)
Donald Trump (Republican)
Jo Jorgensen (Libertarian)
Howie Hawkins (Green)
I voted for someone else not listed here.
I did not vote.

Thinking about your general approach to issues, do you consider yourself to be ________?
Very conservative
Somewhat conservative
Moderate
Somewhat liberal
Very liberal
Not sure

In which state do you live?
▼ Alabama ... District of Columbia

How would describe the area where you live?

- Rural area
- Suburban area
- Urban area

Did you use Google or any other outside sources to answer the questions? Please answer honestly. Your payment does NOT depend on your response to this question.

- No
- Yes

How carefully did you complete this survey? Please answer honestly. Your payment does NOT depend on your response to this question.

- Not at all carefully
- Slightly carefully
- Moderately carefully
- Carefully
- Very carefully

*You might find this information to be useful.*

CDC and FDA have recommended a pause in the use of the Janssen (Johnson & Johnson) COVID-19 vaccine in the United States out of an abundance of caution, effective Tuesday, April 13. Of the 6.8 million Janssen COVID-19 vaccine doses administered in the United States to date, six (6) cases of a type of blood clot called “cerebral venous sinus thrombosis” (CVST) were seen in combination with low levels of blood platelets (thrombocytopenia). All six (6) cases of the “rare and severe” blood clots occurred in women between the ages of 18 and 48, and the symptoms surfaced six (6) to 13 days after the vaccination.

The U.S. Food and Drug Administration (FDA) issued Emergency Use Authorization (EUA) to the single-dose Johnson & Johnson’s Janssen COVID-19 vaccine, to prevent COVID-19 in individuals 18 years of age and older on Feb 27th 2021. Of the 6.8 million Janssen COVID-19 vaccine doses administered in the United States to date, the most common side effects with are usually mild or moderate and get better within 1 or 2 days after vaccination.

*Thank you for taking the survey.*

**eAppdenix 2. Survey Administered to Cohort B**

*Please answer the following questions about your experiences with the COVID-19 pandemic and COVID-19 vaccines.*

Around how often do you look for information about COVID-19 on the internet?

- Never
- Once a month
- 2-3 times per month
- Once a week
- Multiple times per week
- Once a day
- Multiple times per day

Do you ever share information about COVID-19 with your social media network?

- No
- Yes
- I do not have an active account on a social media platform.

Which cable news network do you watch most often?

- CNN
- Fox News
- International news channels (such as BBC, Al Jazeera, or Euronews)
- MSNBC
- I do not watch cable news.

As far as you know, do you have COVID-19 now, or have you had COVID-19 in the past?

- No
- Yes
- I don't know.

As far as you know, has anyone in your immediate social circle, such as close friends or family members, had COVID-19?

- No
- Yes
- I don't know.

Are you currently eligible to get a COVID-19 vaccine where you live?

- No
- Yes

I don't know.

Have you gotten a COVID-19 vaccine?

No

Yes, one dose

Yes, two doses

How likely are you to get vaccinated for COVID-19 when a vaccine is available for you?

I definitely will NOT get vaccinated.

I probably will NOT get vaccinated.

I probably will get vaccinated.

I definitely will get vaccinated.

I am completely undecided about whether I will get vaccinated.

I have already been vaccinated.

Thinking about this in a different way, which of the following statements comes closest to what you are most likely to do when a COVID-19 vaccine is available for you?

I will get vaccinated as soon as possible.

I will wait to see what happens with other people before deciding whether to get vaccinated myself.

I will not get vaccinated, regardless of what happens to other people who get the vaccine.

I have already been vaccinated.

Have you scheduled a COVID-19 vaccine appointment?

Yes, I already got a COVID-19 vaccine.

Yes, I have an upcoming COVID-19 vaccine appointment.

Yes, but my COVID-19 vaccine appointment was cancelled.

No, I don't know how to schedule a COVID-19 vaccine appointment.

No, I am not yet eligible to get a COVID-19 vaccine.

No, I don't plan to schedule a COVID-19 vaccine appointment.

Below are some reasons people may give for why they will NOT get a COVID-19 vaccine. For each one, please indicate whether it definitely applies to you, somewhat applies to you, or does not apply to you as a reason for not getting a COVID-19 vaccine. There is NO right or wrong answer. We are only interested in your personal opinion.

| | Definitely applies to me | Somewhat applies to me | Does not apply to me |
|---|---|---|---|
| I have a health condition that would make it risky for me to get a vaccine. | ◯ | ◯ | ◯ |
| I have a right not to get vaccinated and choose to exercise that right. | ◯ | ◯ | ◯ |
| I do not trust vaccines in general. | ◯ | ◯ | ◯ |
| I do not think the vaccine will be effective in protecting me from COVID-19. | ◯ | ◯ | ◯ |
| I do not think the vaccine will be safe because it could have harmful side effects. | ◯ | ◯ | ◯ |
| I am concerned approval of the vaccine has been rushed, without adequate testing for safety and effectiveness. | ◯ | ◯ | ◯ |
| I do not think it is necessary to get vaccinated because COVID-19 is not as big of a problem as it is being made out to be. | ◯ | ◯ | ◯ |
| I do not think it is necessary to get vaccinated because I take precautions to protect myself, such as wearing a mask, maintaining physical distance, and washing hands often. | ◯ | ◯ | ◯ |
| I do not think I will get very sick if I get COVID-19. | ◯ | ◯ | ◯ |
| I am not able to get a COVID-19 vaccine appointment. | ◯ | ◯ | ◯ |

*Please answer the following two questions just to show that you're paying attention to the survey.*

Please answer “Slightly unlikely” to this question.

- Very unlikely
- Unlikely
- Slightly unlikely
- Slightly likely
- Likely
- Very likely

What color is the sky? Please answer this question incorrectly, on purpose, by choosing “Red” instead of “Blue.”

- Blue
- Green
- Red
- Yellow

*Please read the following text passage about the J&J/Janssen COVID-19 vaccine. After you read the passage, you will be asked some questions. You may take as much time as you would like to read the passage, and you may look back at the passage as you answer the questions.*

***

**Recommendation to Pause Use of Johnson & Johnson’s Janssen COVID-19 Vaccine**

**What you need to know:**

The use of Johnson & Johnson’s Janssen (J&J/Janssen) COVID-19 Vaccine is paused for now. This is because the safety systems that make sure vaccines are safe received a small number of reports of people who got this vaccine experiencing a rare and severe type of blood clot with low platelets. Seek medical care right away if you develop any of the symptoms below. If you have any questions at all, call your doctor, nurse, or clinic.

**J&J/Janssen Vaccine Pause Questions and Answers**

**What if I got the J&J/Janssen COVID-19 Vaccine?**

If you got this vaccine more than three weeks ago, your risk of developing a blood clot with low platelets is very low.

If you got this vaccine within the last three weeks, your risk of developing a blood clot with low platelets is also very low. However, you should be on the lookout for possible symptoms of a blood clot with low platelets:

- severe headache
- backache
- blurred vision
- fainting
- seizures
- severe pain in your abdomen or stomach
- severe pain in your chest
- leg swelling
- shortness of breath
- tiny red spots on the skin (petechiae)
- new or easy bruising or bleeding

**Should I cancel my vaccination appointment?**

If you have an appointment to get the J&J/Janssen COVID-19 Vaccine, please work with your vaccine provider to reschedule your appointment to receive another authorized and recommended COVID-19 vaccine. There are two other COVID-19 vaccines authorized and recommended for use in the United States: Pfizer-BioNTech and Moderna.

**What does a pause mean?**

On April 13, 2021, CDC and the US Food and Drug Administration (FDA) recommended a pause in the use of J&J/Janssen COVID-19 Vaccine. Although the J&J/Janssen vaccine is still authorized for use, CDC and FDA recommend this vaccine not be given to anyone at this time while this safety signal and its possible implications are investigated. The pause will give scientists a chance to review the data and decide if recommendations on this specific vaccine need to change. CDC and FDA will share more information on this situation as soon as possible.

**What do we know?**

Scientists and doctors look constantly and carefully at all reported vaccine side effects.

From their monitoring, they saw a small number of reports of people who got the J&J/Janssen COVID-19 Vaccine developing a rare and severe type of blood clot. This type of blood clot is found in the blood vessels that drain blood from the brain and is combined with low platelets. Platelets help blood clot and stop bleeding.

All of these reports were in women between the ages of 18 and 48, and the problems were found up to 2 weeks after vaccination.

There had been more than 7.5 million doses of the J&J/Janssen COVID-19 Vaccine administered as of the time of the pause in the United States.

**What are we still learning?**

We do not know enough yet to say if the vaccine is related to or caused this health issue. To be extra careful, CDC and FDA recommend that the vaccine not be given until we learn more.

**Why did CDC and FDA recommend a pause?**

CDC and FDA recommended this pause to give the agencies time to communicate with and prepare the healthcare system to recognize and treat patients appropriately, as well as gather more information about this situation. Communication with healthcare providers is also re-emphasizing the importance of reporting and how to report severe events in people who have received this vaccine. This pause also will allow CDC's independent advisory committee, the Advisory Committee on Immunization Practices, to meet, review these reports, and assess their potential significance.

Report any adverse events after vaccination to v-safe and the Vaccine Adverse Event Reporting System.

**What do I need to know about the possible safety issue?**

COVID-19 vaccine safety is a top priority for the federal government, and all reports of health problems following COVID-19 vaccination are taken very seriously.

We know the safety systems in place are working. This potential safety issue was caught early, and this pause reflects the federal government's commitment to transparency as CDC and FDA review these data. COVID-19 vaccines have undergone and will continue to undergo the most intensive safety monitoring in U.S. history.

If you or your patients experience any adverse events after vaccination, report them to v-safe and the Vaccine Adverse Event Reporting System.

**Has this issue been seen with the other COVID-19 vaccines?**

No. As of April 13, 2021, no reports of blood clots with low platelets have been reported among the more than 180 million doses of the Pfizer-BioNTech or Moderna vaccines administered so far.

***

Thinking about the information you just read, how confident are you in the safety of COVID-19 vaccines in general?

Not confident at all
Slightly confident
Somewhat confident
Fairly confident

Completely confident

Thinking about the information you just read, how confident are you in the safety of the J&J/Janssen COVID-19 vaccine in particular?

Not confident at all
Slightly confident
Somewhat confident
Fairly confident
Completely confident

Thinking about the information you just read, how likely would you be to get either the Moderna or Pfizer/BioNTech COVID-19 vaccine?

I definitely would NOT get either the Moderna or Pfizer/BioNTech vaccine.
I probably would NOT get either the Moderna or Pfizer/BioNTech vaccine.
I probably would get either the Moderna or Pfizer/BioNTech vaccine.
I definitely would get either the Moderna or Pfizer/BioNTech vaccine.
I am completely undecided about whether I would get either the Moderna or Pfizer/BioNTech vaccine.
I have already gotten either the Moderna or Pfizer/BioNTech vaccine.
I have already gotten a different COVID-19 vaccine.

*Please answer the same three questions as if we had asked you BEFORE the pause of the J&J/Janssen vaccine was announced.*

Now imagine that we had asked you these three questions BEFORE the pause of the J&J/Janssen vaccine was announced. How would you have answered the first question, which is how confident are you in the safety of COVID-19 vaccines in general?

Not confident at all
Slightly confident
Somewhat confident
Fairly confident
Completely confident

Now imagine that we had asked you these three questions BEFORE the pause of the J&J/Janssen vaccine was announced. How would you have answered the second question, which is how confident are you in the safety of the J&J/Janssen COVID-19 vaccine in particular?

Not confident at all
Slightly confident
Somewhat confident
Fairly confident

Completely confident

Now imagine that we had asked you these three questions BEFORE the pause of the J&J/Janssen vaccine was announced. How would you have answered the third question, which is how likely would you be to get either the Moderna or Pfizer/BioNTech COVID-19 vaccine if available to you?

I definitely would NOT get either the Moderna or Pfizer/BioNTech vaccine.
I probably would NOT get either the Moderna or Pfizer/BioNTech vaccine.
I probably would get either the Moderna or Pfizer/BioNTech vaccine.
I definitely would get either the Moderna or Pfizer/BioNTech vaccine.
I am completely undecided about whether I would get either the Moderna or Pfizer/BioNTech vaccine.
I have already gotten either the Moderna or Pfizer/BioNTech vaccine.
I have already gotten a different COVID-19 vaccine.

*Using a six-point scale, please indicate how strongly you agree with each of the following statements about the passage.*

I think that the information in the passage is accurate and should be trusted.

Strongly disagree
Disagree
Slightly disagree
Slightly agree
Agree
Strongly agree

I think that the information in the passage is based on high-quality evidence.

Strongly disagree
Disagree
Slightly disagree
Slightly agree
Agree
Strongly agree

I think that the writing in the passage is clear and easy to read.

Strongly disagree
Disagree
Slightly disagree
Slightly agree
Agree

Strongly agree

I think that I understand the information in the passage.

Strongly disagree
Disagree
Slightly disagree
Slightly agree
Agree
Strongly agree

I think that I had to put a lot of effort into understanding the information in the passage.

Strongly disagree
Disagree
Slightly disagree
Slightly agree
Agree
Strongly agree

I think that other people would want to read the passage.

Strongly disagree
Disagree
Slightly disagree
Slightly agree
Agree
Strongly agree

I think that I would share the information in the passage with friends on social media.

Strongly disagree
Disagree
Slightly disagree
Slightly agree
Agree
Strongly agree

*Please answer the following questions with your best guess.*

As you read in the passage, more than 7.5 million people have received the J&J/Janssen vaccine in the U.S. *If you had to guess*, about how many of them have developed a rare and severe type of blood clot after being vaccinated?

1 person

10 people
100 people
1,000 people
10,000 people
100,000 people
1,000,000 or more people

As you read in the passage, more than 7.5 million people have received the J&J/Janssen vaccine in the U.S. *If you had to guess*, about how many of them have died from a rare and severe type of blood clot after being vaccinated?

0 people
1 person
10 people
100 people
1,000 people
10,000 people
100,000 people
1,000,000 or more people

*If you had to guess*, how much longer do you think the pause in use of the J&J/Janssen COVID-19 vaccine will last?

1 day
2-6 days
1-2 weeks
2-4 weeks
1-2 months
More than 2 months (but not permanent)
I think the pause will be permanent.

*Thank you for reading the text passage about the J&J/Janssen COVID-19 vaccine and answering our questions.*

*Now please answer a second set of questions based only on the information in the same passage that you just read. As before, you may take as much time as you would like to reread the passage, and you may look back at the passage as you answer the questions.*

Is it currently legal to give the J&J/Janssen vaccine in the U.S.?

No, because the FDA has removed its emergency authorization.
No, because its use has been paused by the FDA and CDC.
Yes, because the pause in use is a recommendation, not a requirement.

The passage does not say.

Why are the CDC and FDA recommending a pause in use of the J&J/Janssen vaccine?

After being approved for emergency use in the U.S., the vaccine was found to be ineffective at preventing COVID-19 infections, hospitalizations, and deaths.

Problems were recently identified with how the original clinical trial was run.

J&J has had trouble making the vaccine, and almost no doses are available now.

A possible safety issue has been identified with the vaccine, and more time is needed to study the issue.

Does the J&J/Janssen COVID-19 vaccine cause blood clots?

No

Yes

More information is needed to know for sure.

A rare and severe type of blood clot has been reported in ________ who received the J&J/Janssen vaccine.

women

men

both men and women

people over the age of 50

On April 1st John got his first shot of the Pfizer/BioNTech vaccine. The day after getting vaccinated, he felt tired and had a mild headache. His second shot is scheduled for this week. Should John cancel the appointment because of safety concerns?

No, because a rare and severe type of blood clot has only been reported in people who received the J&J/Janssen vaccine, not people who received the Moderna or Pfizer/BioNTech vaccines.

No, because blood clots are only caused by the first dose of Pfizer/BioNTech vaccine, not the second.

Yes, because he had a headache after the first dose.

Yes, because possible safety issues with the J&J/Janssen vaccine might apply to all COVID-19 vaccines.

Elizabeth got the the J&J/Janssen COVID-19 six weeks ago. Since then she has not had any side effects from the vaccine or symptoms of COVID-19. In light of the pause, she should ________.

get the Moderna vaccine

get the Pfizer/BioNTech vaccine

ask her doctor for advice about getting a different vaccine

Linda gets the J&J/Janssen vaccine on Monday. On Friday she develops a severe headache. What should she do first?

Seek urgent medical care for a possible blood clot.
Monitor herself for 24 hours and then seek medical care if the headache has not improved.
Do nothing, because headaches are a normal side effect of COVID-19 vaccines.
Report the symptom through v-safe.

Jessie's appointment to get the J&J/Janssen vaccine has been canceled because of the pause. What should they do now?

Work with their vaccine provider to reschedule the appointment and get a different COVID-19 vaccine.
Wait to get vaccinated until the J&J/Janssen vaccine is available again.
Show up for the canceled appointment and ask to get a different vaccine.
Either the first or the second answer is correct.

*Please answer the following questions about your background.*

How old are you?

18-29
30-39
40-49
50-64
65 or older

Which best describes your gender? Please choose as many options as apply.

Female
Male
Non-binary
Transgender
Another option not listed here (please specify):
____________________________________________________

Are you Hispanic or Latino/Latina/Latinx?

Not Hispanic or Latino/Latina/Latinx
Hispanic or Latino/Latina/Latinx

Which best describes your race? Please choose as many options as apply.

American Indian or Alaska Native
Asian

Black or African American
Native Hawaiian or Other Pacific Islander
White
Another option not listed here (please specify):
________________________________________

What is your highest level of formal education?
Some high school or less
High school diploma or equivalent
Some college or associate's degree
Bachelor's degree
Graduate or professional degree

Generally speaking, which of the following best describes your political affiliation?
Strong Republican
Leaning Republican
Leaning Democratic
Strong Democratic
Independent
Another option not listed here (please specify):
________________________________________

Which candidate did you vote for in the 2020 U.S. presidential election?
Joe Biden (Democratic)
Donald Trump (Republican)
Jo Jorgensen (Libertarian)
Howie Hawkins (Green)
I voted for someone else not listed here.
I did not vote.

Thinking about your general approach to issues, do you consider yourself to be ________?
Very conservative
Somewhat conservative
Moderate
Somewhat liberal
Very liberal
Not sure

In which state do you live?

▼ Alabama ... District of Columbia

How would describe the area where you live?

Rural area
Suburban area
Urban area

Did you use Google or any other outside sources to answer the questions? Please answer honestly. Your payment does NOT depend on your response to this question.

No
Yes

How carefully did you complete this survey? Please answer honestly. Your payment does NOT depend on your response to this question.

Not at all carefully
Slightly carefully
Moderately carefully
Carefully
Very carefully

*You might find this information to be useful.*

CDC and FDA have recommended a pause in the use of the Janssen (Johnson & Johnson) COVID-19 vaccine in the United States out of an abundance of caution, effective Tuesday, April 13.

Scientists and doctors look constantly and carefully at all reported vaccine side effects.

About 7 million Janssen COVID-19 vaccine doses administered in the United States to date, six (6) cases of a rare and severe type of blood clot called "cerebral venous sinus thrombosis" (CVST) were seen in combination with low levels of blood platelets (thrombocytopenia). This type of blood clot is found in the blood vessels that drain blood from the brain and is combined with low platelets. Platelets help blood clot and stop bleeding.

All six (6) cases of the "rare and severe" blood clots occurred in women between the ages of 18 and 48, and the problems were found up to 2 weeks after vaccination.

When these specific types of blood clots are observed following J&J COVID-19 vaccination, treatment is different from the treatment that might typically be administered for blood clots. The purpose of this Health Alert is, in part, to ensure that the healthcare provider community is aware of the potential for these adverse events and can provide proper management due to the unique treatment required with this type of blood clot.

The U.S. Food and Drug Administration (FDA) issued Emergency Use Authorization (EUA) to the single-dose Johnson & Johnson's Janssen COVID-19 vaccine, to prevent COVID-19 in individuals 18 years of age and older on Feb 27th 2021. Of all the Janssen COVID-19 vaccine doses administered in the United States to date, the most common side effects with are usually mild or moderate and get better within 1 or 2 days after vaccination.

*Thank you for taking the survey.*

### eTable 1. Full Responses to Comprehension Questions

| Question, Answer Choices[a] | Cohort A (N = 271) No. (%) | Cohort B (N = 286) No. (%) |
|---|---|---|
| Why are the CDC and FDA recommending a pause in use of the J&J/Janssen vaccine? | | |
| (1) A possible safety issue has been identified with the vaccine, and more time is needed to study the issue. | 259 (95.6) | 280 (97.9) |
| (2) After being approved for emergency use in the U.S., the vaccine was found to be ineffective at preventing COVID-19 infections, hospitalizations, and deaths. | 4 (1.5) | 2 (0.70) |
| (3) Problems were recently identified with how the original clinical trial was run. | 4 (1.5) | 4 (1.4) |
| (4) J&J has had trouble making the vaccine, and almost no doses are available now. | 4 (1.5) | 0 (0) |
| Does the J&J/Janssen COVID-19 vaccine cause blood clots? | | |
| (1) More information is needed to know for sure. | 196 (72.3) | 215 (75.2) |
| (2) No | 3 (1.1) | 4 (1.4) |
| (3) Yes | 72 (26.6) | 67 (23.4) |
| A rare and severe type of blood clot has been reported in ________ who received the J&J/Janssen vaccine. | | |
| (1) women | 234 (86.3) | 248 (86.7) |
| (2) men | 1 (0.4) | 0 (0) |
| (3) both men and women | 28 (10.3) | 36 (12.6) |
| (4) people over the age of 50 | 8 (3.0) | 2 (0.7) |
| On April 1st John got his first shot of the Pfizer/BioNTech vaccine. The day after getting vaccinated, he felt tired and had a mild headache. His second shot is scheduled for this week. Should John cancel the appointment because of safety concerns? | | |
| (1) No, because a rare and severe type of blood clot has only been reported in people who received the J&J/Janssen vaccine, not people who received the Moderna or Pfizer/BioNTech vaccines. | 214 (79.0) | 250 (87.0) |
| (2) No, because blood clots are only caused by the first dose of Pfizer/BioNTech vaccine, not the second. | 5 (1.8) | 3 (1.0) |
| (3) Yes, because he had a headache after the first dose. | 18 (6.6) | 14 (4.9) |
| (4) Yes, because possible safety issues with the J&J/Janssen vaccine might apply to all COVID-19 vaccines. | 34 (12.5) | 19 (6.6) |
| Elizabeth got the the J&J/Janssen COVID-19 six weeks ago. Since then she has not had any side effects from the vaccine or symptoms of COVID-19. In light of the pause, she should ________. | | |
| (1) do nothing different | 164 (60.5) | 189 (66.1) |
| (2) get the Moderna vaccine | 1 (0.4) | 1 (0.3) |
| (3) get the Pfizer/BioNTech vaccine | 8 (3.0) | 8 (2.8) |
| (4) ask her doctor for advice about getting a different vaccine | 98 (36.2) | 88 (30.8) |
| Linda gets the J&J/Janssen vaccine on Monday. On Friday she develops a severe headache. What should she do first? | | |
| (1) Seek urgent medical care for a possible blood clot. | 180 (66.4) | 121 (42.3) |
| (2) Monitor herself for 24 hours and then seek medical care if the headache has not improved. | 62 (22.9) | 87 (30.4) |
| (3) Do nothing, because headaches are a normal side effect of COVID-19 vaccines. | 8 (3.0) | 4 (1.4) |
| (4) Report the symptom through v-safe. | 28 (10.3) | 74 (25.9) |

| | | |
|---|---|---|
| Jessie's appointment to get the J&J/Janssen vaccine has been canceled because of the pause. What should they do now?[b] | | |
| (1) Work with their vaccine provider to reschedule the appointment and get a different COVID-19 vaccine. | 192 (70.8) | 229 (80.1) |
| (2) Wait to get vaccinated until the J&J/Janssen vaccine is available again. | 10 (3.7) | 6 (2.1) |
| (3) Show up for the canceled appointment and ask to get a different vaccine. | 9 (3.3) | 4 (1.4) |
| (4) Either the first or the second answer is correct. | 10 (3.7) | 47 (16.4) |

[a]Unless otherwise indicated, answer choices were presented to the participants in a random order. For clarity the correct answer is always listed first in the table.
[b]Order of answer choices was not randomized.

### eTable 2. Summary of Ordinal Logistic Regression Analysis for Cohort A

| **Feature** | **Odds Ratio** | **95% CI** | ***P*** |
|---|---|---|---|
| Age | | | |
| ≥50 | 0.94 | 0.64-1.39 | 0.77 |
| Gender | | | |
| Female | 1.18 | 0.92-1.53 | 0.20 |
| Ethnicity | | | |
| Hispanic or Latinx | 0.83 | 0.59-1.17 | 0.30 |
| Race | | | |
| Asian | 1.13 | 0.72-1.77 | 0.61 |
| Black or African American | 0.78 | 0.56-1.08 | 0.13 |
| Other | 1.16 | 0.77-1.75 | 0.48 |
| Educational attainment | | | |
| High school diploma or less | 0.66 | 0.48-0.89 | 0.007 |
| Political partisanship | | | |
| Republican | 1.25 | 0.93-1.69 | 0.14 |
| Geography | | | |
| Rural area | 1.12 | 0.77-1.63 | 0.54 |
| Intention to receive vaccine | | | |
| Definitely will not | 0.61 | 0.45-0.82 | 0.001 |

### eTable 3. Summary of Ordinal Logistic Regression Analysis for Cohort B

| **Feature** | **Odds Ratio** | **95% CI** | ***P*** |
| --- | --- | --- | --- |
| Age | | | |
| ≥50 | 1.26 | 0.83-1.91 | 0.28 |
| Gender | | | |
| Female | 1.33 | 1.03-1.71 | 0.027 |
| Ethnicity | | | |
| Hispanic or Latinx | 1.09 | 0.71-1.65 | 0.73 |
| Race | | | |
| Asian | 0.85 | 0.62-1.15 | 0.15 |
| Black or African American | 0.46 | 0.31-0.68 | <0.001 |
| Other | 0.85 | 0.53-1.35 | 0.41 |
| Educational attainment | | | |
| High school diploma or less | 0.79 | 0.57-1.09 | 0.15 |
| Political partisanship | | | |
| Republican | 0.85 | 0.59-1.23 | 0.56 |
| Geography | | | |
| Rural area | 0.89 | 0.60-1.32 | 0.39 |
| Intention to receive vaccine | | | |
| Definitely will not | 0.48 | 0.31-0.74 | 0.001 |

**eReferences**


**1.** Ndugga N, Pham O, Hill L, Artiga S. Latest data on COVID-19 vaccinations race/ethnicity. Published May 26, 2021. Accessed June 6, 2021. https://www.kff.org/coronavirus-covid-19/issue-brief/latest-data-on-covid-19-vaccinations-race-ethnicity/

**2.** Grumbach K, Judson T, Desai M, Jain V, Lidan C, Doernberg SB, Holubar M. Association of race/ethnicity with likeliness of COVID-19 vaccine uptake among health workers and the general population in the San Francisco Bay Area. *JAMA Intern Med*. Published online March 30, 2021. doi:10.1001/jamainternmed.2021.1445

**3.** Persad G, Emanuel EJ, Sangenito S, Glickman A, Phillips S, Largent EA. Public perspectives on COVID-19 vaccine prioritization. *JAMA Netw Open* 2021;4(4):e217943. doi:10.1001/jamanetworkopen.2021.7943

**4.** Marshall MN. Sampling for qualitative research. *Fam Pract*. 1996;13(6):522-525. doi:10.1093/fampra/13.6.522

**5.** Kerr JR, Freeman ALJ, Marteau TM, van der Linden S. Effect of information about COVID-19 vaccine effectiveness and side effects on behavioural intentions: Two online experiments. *Vaccines* 2021:9(4):379. doi:10.3390/vaccines9040379

**6.** Graham MH, Coppock A. Asking about attitude change. Published February 2, 2021. Accessed June 6, 2021. https://m-graham.com/papers/GrahamCoppock_aaac.pdf

**7.** Graham M, Coppock A. How to use the counterfactual polling format to ask about attitude change. Published April 15, 2021. Accessed June 6, 2021. https://alexandercoppock.com/subpages/counterfactual_format.html
